# Forecasts of weekly incident and cumulative COVID-19 mortality in the United States: A comparison of combining methods

**DOI:** 10.1101/2021.07.11.21260318

**Authors:** Kathryn S. Taylor, James W. Taylor

**Affiliations:** Nuffield Department of Primary Care Health Sciences, University of Oxford, Oxford, United Kingdom; Saïd Business School, University of Oxford, Oxford, United Kingdom

## Abstract

**Background:** Forecasting models have played a pivotal role in health policy decision making during the coronavirus disease-2019 (COVID-19) pandemic. A combined forecast from multiple models will be typically more accurate than an individual forecast, but there are few examples of studies of combined forecasts of COVID-19 data, focusing mainly on simple mean and median ‘ensembles’ and involving short forecast evaluation periods. We aimed to investigate the accuracy of different ways of combining probabilistic forecasts of weekly COVID-19 mortality data, including two weighted methods that we developed previously, on an extended dataset and new dataset, and evaluate over a period of 52 weeks.

**Methods:** We considered 95% interval and point forecasts of weekly incident and cumulative COVID-19 mortalities between 16 May 2020 and 8 May 2021 in multiple locations in the United States. We compared the accuracy of simple and more complex combining methods, as well as individual models.

**Results:** The average of the forecasts from the individual models was consistently more accurate than the average performance of these models (the mean combination), which provides a fundamental motivation for combining. Weighted combining performed well for both incident and cumulative mortalities, and for both interval and point forecasting. Our inverse score with tuning method was the most accurate overall. The median combination was a leading method in the last quarter for both mortalities, and it was consistently more accurate than the mean combination for point forecasting. For interval forecasts of cumulative mortality, the mean performed better than the median. The best performance of the leading individual model was in point forecasting.

**Conclusions:** Combining forecasts can improve the contribution of probabilistic forecasting to health policy decision making during epidemics, and, when there are sufficient historical data on forecast accuracy, weighted combining provides the most accurate method.

## Introduction

The coronavirus disease-2019 (COVID-19) pandemic has overwhelmed health services and caused excess death rates, prompting governments to impose extreme restrictions in attempts to control the spread of the virus [1–3]. These interventions have resulted in multiple economic, health and societal problems [4, 5]. This has generated intense debate among experts about the best way forward[6]. Governments and their advisors have relied upon forecasts from models of the numbers of COVID-19 cases, hospitalisations and deaths to help decide what actions to take [7]. Using models to lead health policy has been controversial, but it is recognised that modelling is potentially valuable when used appropriately [1, 8–10]. Numerous models have been developed to forecast different COVID-19 data, e.g. [11–13].

Models should provide probabilistic forecasts, as point forecasts are inherently uncertain [9, 14]. A 95% interval forecast is a common and useful form of probabilistic forecast [15] [16]. Models may be constructed for prediction or scenario analysis. Prediction models forecast the most likely outcome in the current circumstances. Multiple models may reflect different approaches to answering the same question [11], and conflicting forecasts may arise. Rather than asking which is the best model [17], a forecast combination can be used, such as the mean, which is often used and hard to beat [18, 19]. Forecast combining harnesses the ‘wisdom of the crowd’ [20], that is, produces a collective forecast from multiple models that is typically more accurate than forecasts from individual models. Combining pragmatically synthesises information underlying different prediction methods, diversifying the risk inherent in relying on an individual model, and it can offset statistical bias, potentially cancelling out overestimation and underestimation [21]. These advantages are well-established in many applications outside health care [22–25]. This has encouraged the more recent applications of combining in infectious disease prediction [14, 26–29], including online platforms that present visualisations of simple combined probabilistic forecasts of COVID-19 data from the U.S, reported by the Centre for Disease Control and Prevention (CDC), and from Europe, reported by the European Centre for Disease and Control (EDCD). Other examples or combined probabilistic forecasts are in vaccine trial planning [30] and diagnosing disease [31]. These examples have focused on simple mean and median ‘ensembles’ and, in the case of prediction of COVID-19 data, published studies have involved short forecast evaluation periods, which rules out the consideration of more sophisticated methods, such as those weighted by historical accuracy.

By comparing the accuracy of different combining methods over longer forecast evaluation periods compared to other studies, our broad aims were to: (a) investigate whether combining methods, involving weights determined by prior forecast accuracy or different ways of excluding outliers, are more accurate than simple methods of combining, and (b) establish the relative accuracy of the two simple methods. Previously, we reported several new weighted methods, in a comparison of combining methods applied to probabilistic predictions of weekly cumulative COVID-19 mortality in U.S. locations over the 40-week period up to 23 January 2021 [32]. We found that weighted methods were the most accurate overall and the mean generally outperformed the median except in the first ten weeks. In this paper, we test further by comparing the combining methods on an extended dataset of cumulative mortality and a new dataset of incident mortality, over a longer period of 52 weeks. Here, we also include individual models in the comparison, and explore the impact of reporting delays of death counts on forecast accuracy.

## Materials and methods

### Data sources

Forecasts of weekly incident and cumulative COVID-19 mortalities were downloaded from the COVID-19 Forecast Hub [26, 33], which is an ongoing collaboration between the U.S. Centers for Disease Control and Prevention (CDC), with forecasting teams from academia, industry and government-affiliated groups [33]. Teams are invited to submit forecasts for 1-to 4-week horizons, in the form of estimates of quantiles corresponding to 23 probability points along the probability distribution, as well as point forecasts. We considered 95% interval forecasts (bound by the 2.5% and 97.5% quantiles), and median point forecasts (the 50% quantile). The numbers of actual cumulative COVID-19 deaths each week were also provided by the Hub. Their reference data source is the Centre for Systems Science and Engineering (CSSE) at John Hopkins University [33].

### Analysis dataset

We included forecasts projected from forecast origins between the weeks ending 16 May 2020 (Epidemic Week 21) and 8 May 2021 (Week 72), and we considered actual weekly COVID-19 mortality up to the week ending 15 May 2021 (Week 73). We studied forecasts of COVID-19 mortality for the U.S. as a whole and 51 U.S. jurisdictions, including the 50 states and the District of Columbia. For simplicity, we refer to 51 states.

The Hub carries out screening tests for inclusion in their ‘ensemble’ forecast combination. We included forecasts that passed the Hub’s screening tests, and forecasts that were not submitted in time for screening. For any given week, we excluded a model for which forecasts for all 23 quantiles and for all four forecast horizons were not provided. The Hub also excludes outliers. We did not exclude outliers, primarily because the actual number of COVID-19 deaths in previous weeks may have been updated, and therefore the assessment of outliers in the past would not be consistent with our retrospective assessments of outliers at the end of the dataset.

### Evaluating the forecasts

The accuracy of the 95% interval forecasts was evaluated in terms of calibration and the interval score. Calibration was assessed by the percentage of actual deaths that fell within the bounds of the interval forecasts, with the ideal being 95%. The interval score was calculated by the following expression [34, 35]:

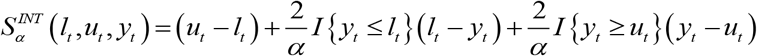

where *l*_*t*_ is the interval’s lower bound, *u*_*t*_ is its upper bound, *y*_*t*_ is the observation in period *t, I* is the indicator function (1 if the condition is true and 0 otherwise), and, for a 95% interval, *α*=5%. Lower values of the interval score reflect greater interval forecast accuracy. This score are useful for comparing methods, and although its unit is deaths, the score is not interpretable. The accuracy of the point forecasts were evaluated using the absolute value of the forecast errors. Averaging each of these two scores across weeks provided two measures of forecast accuracy – the mean interval score (MIS) and the mean absolute error (MAE).

We also averaged across horizons, for conciseness, and because we had a relatively short analysis period, which is a particular problem when evaluating forecasts of extreme quantiles. To show the consistency across horizons, we present results by horizon for interval forecasts. We adapted the Diebold-Mariano statistical test [36] to test across multiple series, for each prediction horizon. To summarise results averaged across the four horizons, we applied the statistical test proposed by Koning et al. [37], which, for each method, compares the rank, averaged across multiple series, with the corresponding average rank of the most accurate method. Statistical testing was based on a 5% significance level.

We evaluated the effects of reporting delays on forecast accuracy by analysing files of actual death counts at 13 different time points between Weeks 26 and 73. Updates, typically increases in death counts, result in forecasting models underestimating, and when updates are backdated, this leads to forecasting methods being penalised in retrospective analyses of forecast accuracy. Analysis was carried out using GAUSS software.

### Forecast combining methods

The comparison included several interval combining methods that do not rely on the availability of records of past accuracy (Fig 1). Combining is applied to each interval bound separately.

**Fig 1.**
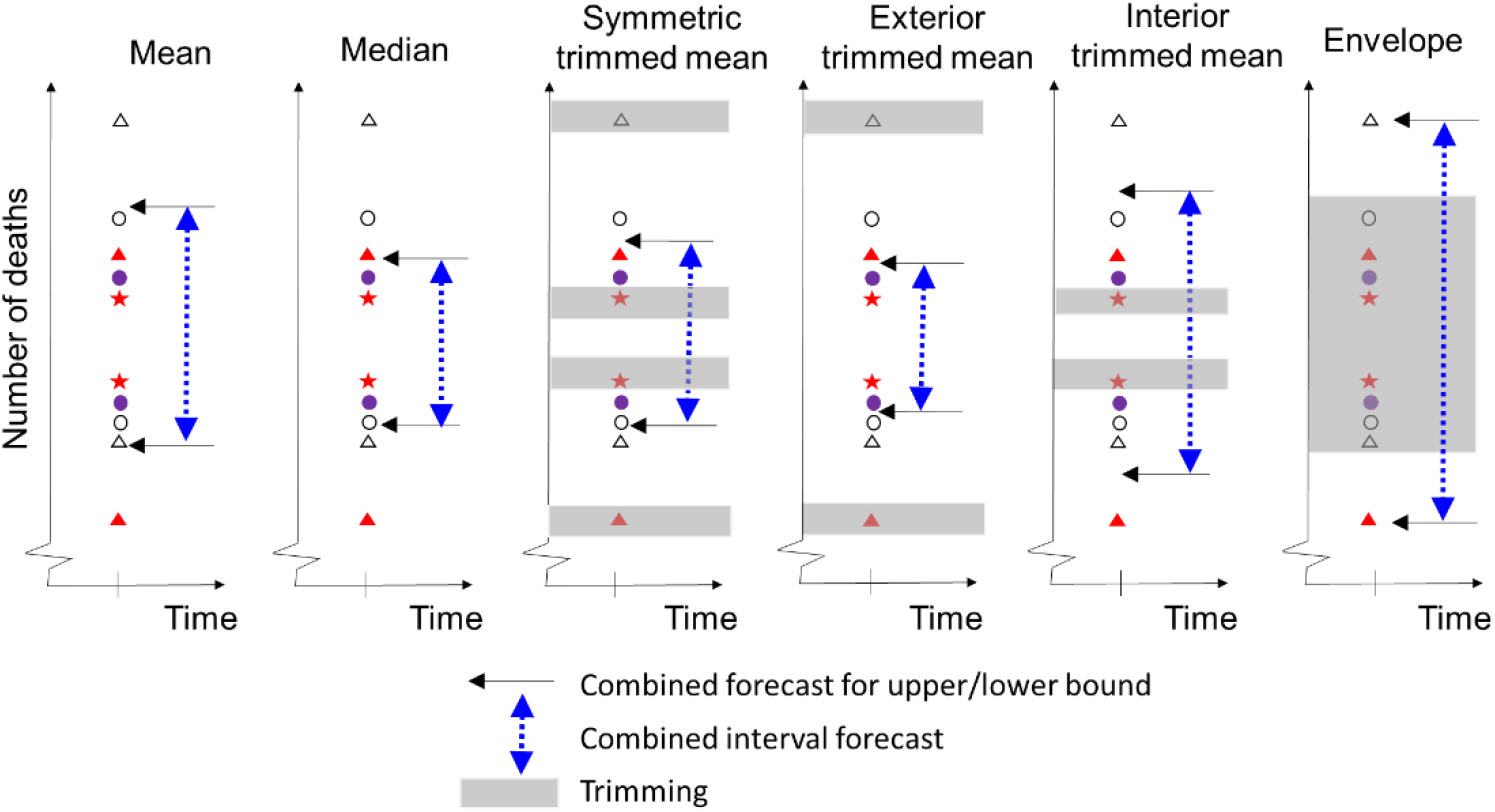
Illustration of interval forecast combining methods that do not rely on past historical accuracy. Each pair of shapes represents an interval forecast produced by an individual model.

These methods include the well-established *mean* and *median* combinations [38–40] and more novel trimming methods, which exclude a particular percentage of forecasts, and then average the remaining forecasts of each bound [41]. Trimming is carried out to either deal with outliers, overconfidence or underconfidence. Trimming methods included *symmetric trimming, exterior trimming, interior trimming* and the *envelope* method. We also included: the COVID-19 Hub’s *ensemble* forecast, which was initially the mean combination of the forecasts that they considered eligible, and later it became the median; the best method at each week, the *previous best* [42]; and, our two weighted combinations, with weights based on the inverse of the MIS [32], the *inverse score* and *inverse score with tuning*. Further details are provided in S1 File.

We divided the 52 weeks into an expanding in-sample period, starting with length 13 weeks, and a 39-week out-of-sample period. For each location and trimming method, we optimised the trimming percentage, by finding the value that minimised the MIS averaged over all four horizons and all periods up to and including the current forecast origin (the in-sample period). The same approach was applied to optimise the tuning parameter. For the inverse score methods, the scores were calculated in-sample.

For the point forecasts, we considered analogous combining methods to those for the interval forecasts, with the exception of the interior and exterior trimming means and envelope combining methods, which have no analogy for combining point forecasts. We included individual models for which forecasts were available for all 52 locations and all 39 out-of-sample periods.

## Results

### Forecasting models

Our analysis included forecasts from 53 forecasting models and the Hub’s ensemble model (S1 Table). In the early weeks of our dataset, the majority were susceptible-exposed-infected-removed (SEIR) compartmental models, but as the weeks passed, other model types became more common (Fig 2). These involved neural networks, agent-based and time series modelling, and curve fitting techniques.

**Fig 2.** Number and types of models at each forecast origin for each mortality dataset.

The timeline of forecasts from each model illustrates the extent of missing data across the 52 locations, including the frequent ‘entry and exit’ of forecasting teams (Fig 3). The extent of missing data was such that imputation was impractical. In comparing forecast accuracy results, we were able to include six individual models for forecasts of incident mortality (numbered 1, 14, 20, 21, 22 and 33, as shown in Fig 3), and two models for cumulative mortality (21 and 33), because these models were available for all locations and weeks in the out-of-sample period.

**Fig 3.**
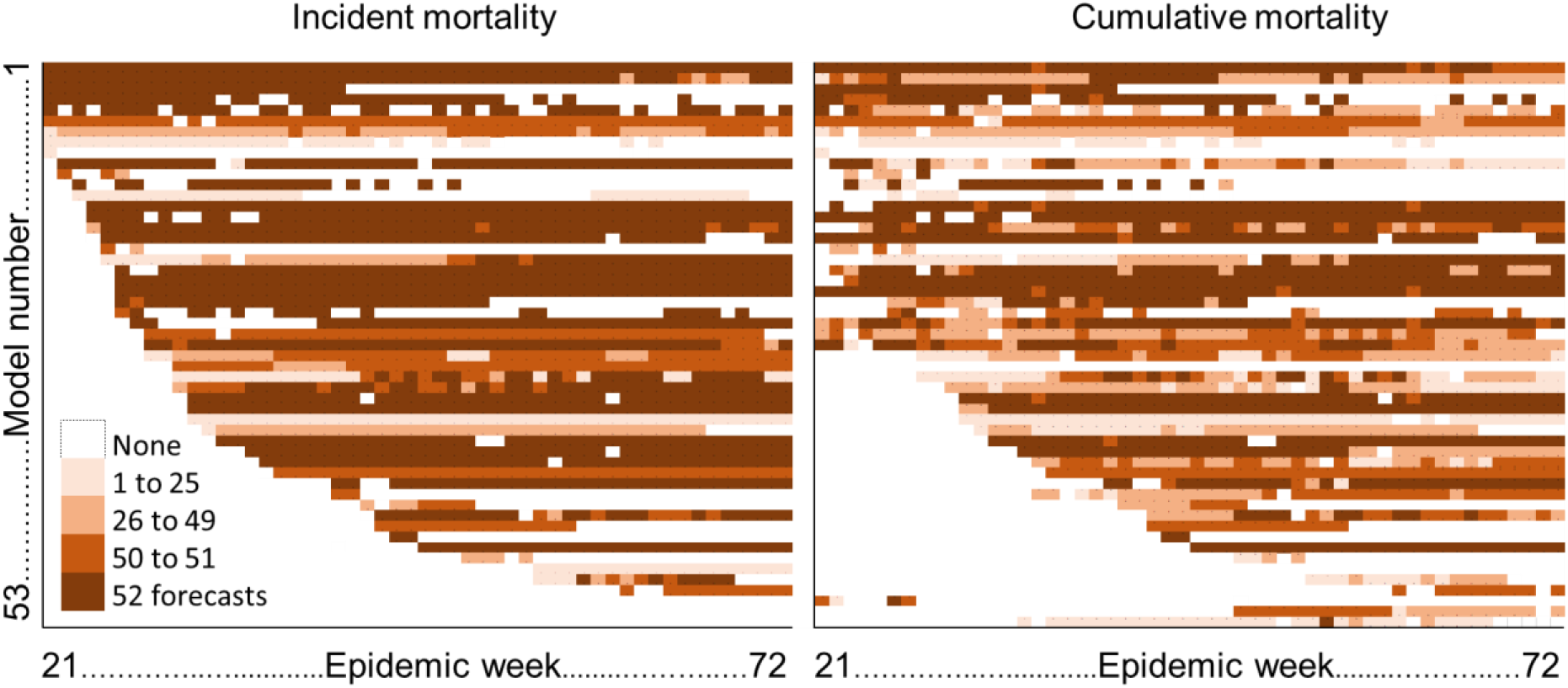
Data availability.

### Performance of methods overall

We present results averaged across all 52 locations, for the whole U.S., and as the unit of scores was deaths, to avoid scores for some locations dominating, we also present results averaged for three categories: high, medium and low mortality states. The categorisation was in accordance with the number of cumulative COVID-19 deaths at the end of Week 73. Tables 1 and 2 present the MIS and mean ranks for 95% interval forecasts for the 32-week out-of-sample period for incident and cumulative mortality, respectively. Tables 3 and 4 present the corresponding MAE results for point forecasts.

**Table 1.** For 95% interval forecasts of incident mortality, mean interval scores and mean ranks for the 39-week out-of-sample period.

| Method | Mean interval score |  |  |  |  | Mean rank |  |  |  |  |
| --- | --- | --- | --- | --- | --- | --- | --- | --- | --- | --- |
|  | All | U.S. | High | Med | Low | All | U.S. | High | Med | Low |
| Mean | 900 | 9799 | 1631 | 443 | 104 | 5.8 | 4 | 7.4 | 5.1 | 5.0 |
| Median | 778 | 11239 | 1172 | 435 | 110 | 5.8 | 9 | 6.4 | 5.0 | 5.9 |
| Ensemble | 794 | 11176 | 1211 | 448 | 112 | 6.7 | 8 | 7.1 | 6.4 | 6.6 |
| Sym trim | 878 | 10445 | 1515 | 446 | 109 | 6 | 6 | 6.3 | 5.8 | 5.8 |
| Exterior trim | 996 | 12447 | 1716 | 480 | 118 | 8.6<sup>†</sup> | 11 | 9.4 | 7.9 | 8.5 |
| Interior trim | 895 | 9969 | 1625 | 429 | 97* | 4.8 | 5 | 7.0 | 4.1* | 3.3 |
| Envelope | 5284 | 64944 | 11037 | 1004 | 300 | 15<sup>†</sup> | 16 | 14.8<sup>†</sup> | 14.6<sup>†</sup> | 15.5<sup>†</sup> |
| Previous best | 877 | 11779 | 1217 | 637 | 135 | 9.8<sup>†</sup> | 10 | 8.7 | 10.9<sup>†</sup> | 9.9<sup>†</sup> |
| Inv score | 770 | 9130 | 1296 | 425* | 98 | 3.9* | 3 | 4.5* | 4.1* | 3.2* |
| Inv score tuning | 677* | 8939 | 1000 | 441 | 104 | 5.2 | 2 | 4.6 | 5.8 | 5.4 |
| Model 1 | 729 | 11174 | 948* | 512 | 114 | 6.3 | 7 | 5.3 | 7.9 | 5.7 |
| Model 14 | 1918 | 30502 | 2936 | 909 | 227 | 13.7<sup>†</sup> | 13 | 13.4<sup>†</sup> | 13.5<sup>†</sup> | 14.2<sup>†</sup> |
| Model 20 | 1957 | 33948 | 2808 | 933 | 248 | 13.5<sup>†</sup> | 14 | 13.4<sup>†</sup> | 13.6<sup>†</sup> | 13.5<sup>†</sup> |
| Model 21 | 2004 | 35399 | 2846 | 949 | 254 | 14.3<sup>†</sup> | 15 | 13.9<sup>†</sup> | 14.5<sup>†</sup> | 14.3<sup>†</sup> |
| Model 22 | 1014 | 19145 | 1312 | 535 | 130 | 9.2<sup>†</sup> | 12 | 8.9 | 9.1 | 9.5<sup>†</sup> |
| Model 33 | 748 | 8681* | 1139 | 499 | 139 | 7.3<sup>†</sup> | 1* | 5.1 | 7.5 | 9.8<sup>†</sup> |
| Model average | 2222 | 37870 | 3315 | 995 | 257 |  |  |  |  |  |

\* Best method in each column; <sup>†</sup>method is significantly worse than method with best mean rank.

**Table 2.** For 95% interval forecasts of cumulative mortality, mean interval scores and mean ranks for the 39-week out-of-sample period.

| Method | Mean interval score |  |  |  |  | Mean rank |  |  |  |  |
| --- | --- | --- | --- | --- | --- | --- | --- | --- | --- | --- |
|  | All | U.S. | High | Med | Low | All | U.S. | High | Med | Low |
| Mean | 3529 | 48497 | 6761 | 925 | 256 | 5.7 | 6 | 5.9 | 4.9 | 6.2 |
| Median | 3700 | 56185 | 6769 | 934 | 310 | 6.1 | 11 | 7.2 | 5.6 | 5.3 |
| Ensemble | 3588 | 50481 | 6747 | 941 | 318 | 6.5 | 8 | 7.1 | 6.1 | 6.2 |
| Sym trim | 3671 | 52563 | 6956 | 924 | 257 | 5.7 | 9 | 6.6 | 5.2 | 4.9 |
| Exterior trim | 3737 | 53839 | 7006 | 985 | 271 | 7.6<sup>†</sup> | 10 | 8.1 | 7.2 | 7.3 |
| Interior trim | 3883 | 36142* | 8623 | 921 | 208* | 4.8 | 1* | 5.0 | 4.7* | 4.8 |
| Envelope | 6545 | 103456 | 11304 | 1968 | 663 | 11.7<sup>†</sup> | 12 | 11.4<sup>†</sup> | 11.7<sup>†</sup> | 12.1<sup>†</sup> |
| Previous best | 3531 | 49146 | 6371 | 1235 | 303 | 8.9<sup>†</sup> | 7 | 7.7 | 10.2<sup>†</sup> | 9.1<sup>†</sup> |
| Inv score | 3296 | 40479 | 6561 | 920* | 221 | 4.2* | 2 | 4.5 | 5.2 | 3.1* |
| Inv score tuning | 3100* | 40758 | 5918* | 948 | 219 | 4.8 | 3 | 4.2* | 6.1 | 4.4 |
| Model 21 | 7247 | 136492 | 11276 | 2202 | 658 | 12.2<sup>†</sup> | 13 | 12.4<sup>†</sup> | 12.1<sup>†</sup> | 12.2<sup>†</sup> |
| Model 33 | 3457 | 44297 | 6552 | 1068 | 348 | 7.4<sup>†</sup> | 4 | 5.3 | 8.0 | 9.2<sup>†</sup> |
| Model average | 6486 | 114911 | 10333 | 2114 | 633 |  |  |  |  |  |

\*Best method in each column; <sup>†</sup>method is significantly worse than method with best mean rank.

**Table 3.**
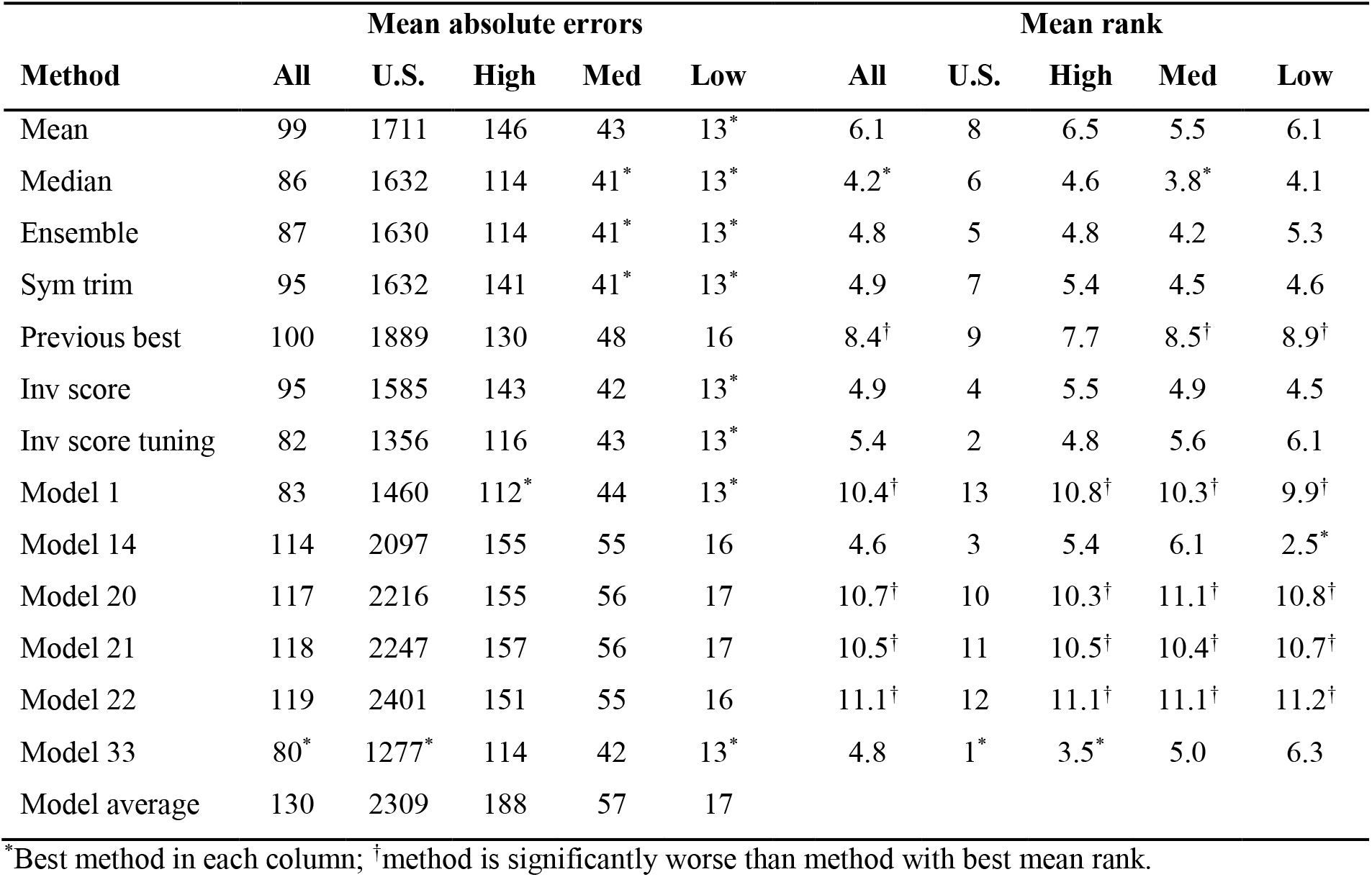
For point forecasts of incident mortality, mean absolute errors and mean ranks for the 39-week out-of-sample period.

**Table 4.** For point forecasts of cumulative mortality, mean absolute errors and mean ranks for the 39-week out-of-sample period

| Method | Mean absolute errors |  |  |  |  | Mean rank |  |  |  |  |
| --- | --- | --- | --- | --- | --- | --- | --- | --- | --- | --- |
|  | All | U.S. | High | Med | Low | All | U.S. | High | Med | Low |
| Mean | 270 | 4754 | 430 | 87 | 30 | 5.5 | 8 | 5.6 | 5.2 | 5.4 |
| Median | 240 | 4438 | 359 | 84* | 30 | 4.4 | 5 | 5.1 | 3.8* | 4.3 |
| Ensemble | 238 | 4373 | 357 | 85 | 30 | 4.8 | 4 | 4.5 | 4.3 | 5.8 |
| Sym trim | 244 | 4639 | 359 | 84* | 29* | 4.5 | 7 | 5.5 | 4.3 | 3.5* |
| Previous best | 253 | 3976 | 398 | 108 | 32 | 7.5<sup>†</sup> | 2 | 6.7<sup>†</sup> | 8.6<sup>†</sup> | 7.4<sup>†</sup> |
| Inv score | 262 | 4576 | 417 | 87 | 29* | 4.8 | 6 | 5.2 | 5.0 | 4.1 |
| Inv score tuning | 236 | 4291 | 353* | 86 | 29* | 4.3* | 3 | 4.4 | 4.6 | 4.0 |
| Model 21 | 346 | 6914 | 480 | 128 | 42 | 9.6<sup>†</sup> | 10 | 9.6<sup>†</sup> | 9.5<sup>†</sup> | 9.7<sup>†</sup> |
| Model 33 | 231* | 3910* | 360 | 88 | 30 | 4.7 | 1* | 3.0* | 4.8 | 6.4 |
| Model average | 338 | 6102 | 515 | 120 | 38 |  |  |  |  |  |

\*Best method in each column; <sup>†</sup>method is significantly worse than method with best mean rank.

Considering methods that performed well in terms of either being the leading method or competitive against the leading method, according to the scores and mean ranks, inverse score combining performed best overall, and benefitted from tuning (Tables 1 to 4). The poorest results were produced by the envelope method (Tables 1 to 4). The interior trimmed mean performed well for interval forecasting for low and medium mortality states, producing the most accurate interval forecasts for the low mortality category for both the incident and cumulative mortality data (Tables 1 and 2). The leading individual model (Model 33) performed well in forecasting the numbers of deaths for all series, the U.S. as a whole, and for high mortality states (Tables 1 to 4). Its performance was stronger in point forecasting than interval forecasting, and it was more accurate in forecasting incident deaths than cumulative deaths (Tables 1 to 4).

The final row of Tables 1 to 4 shows that the average score of all the individual methods was substantially worse than the performance of the mean combining method (the average performance was much worse than the performance of the average).

Comparing the two simple combining methods, for interval forecasts of incident deaths, the median was more accurate than the mean for all series and for high mortality states, whilst the performance was similar for medium and low mortality states (Table 1). For interval forecasts of cumulative mortality, the mean was the more accurate method across all categories (Table 2). The median was consistently the more accurate for point forecasts of both incident and cumulative mortality (Tables 3 and 4).

The methods that performed poorly were identified as being statistically significantly worse than the best methods. Tables 1 to 4 report results averaged across the four forecast horizons (1 to 4 weeks ahead). We found similar relative performances of the methods when looking at each forecast horizon separately for both forecasts of incident mortality (S2 Table) and cumulative mortality (S3 Table).

### Changes in performance over time

For the first 13-week period, results were not obtainable for methods for which a previous in-sample period was needed to estimate parameters. The leading methods in the second and third quarters for forecasts of incident mortality were the inverse score methods (S4 Table), and for cumulative mortality, the inverse score methods and the interior trimmed mean (S5 Table). For forecasts of both mortalities, the leading methods in the most recent quarter were the median-based methods (median and ensemble). However, for both incident and cumulative mortalities, the mean was more accurate than the median in the second and third quarters.

### Impact of reporting delays on performance

We observed updates to historical death counts in 14 locations. Updates were particularly noticeable in six states (Fig 4).

**Fig 4.**
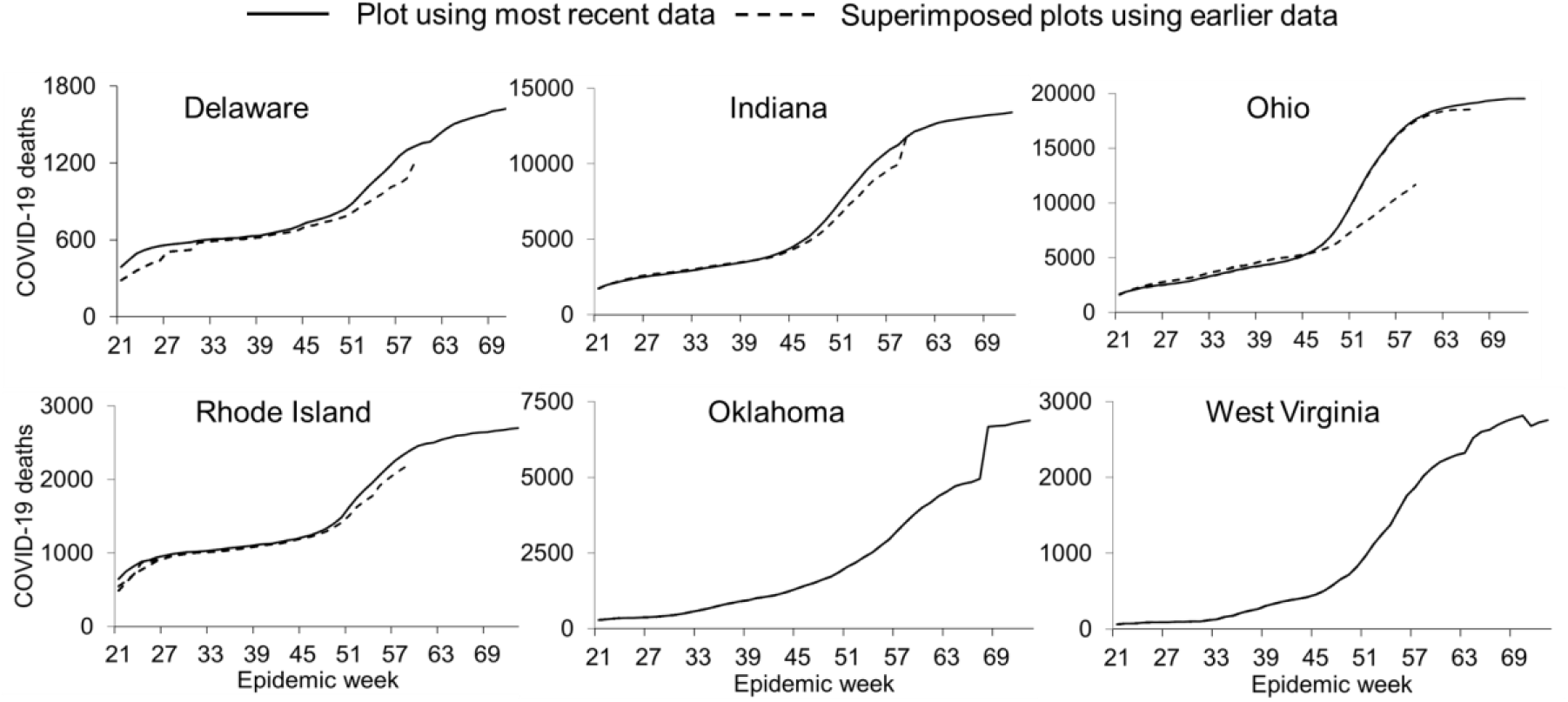
Numbers of reported COVID-19 deaths in states where there were noticeable effects of reporting delays. Shows superimposed plots using data files at Weeks 26, 30, 32, 34, 37, 41, 43, 45, 47, 54, 59, 66 and 73.

Fig 5 presents the MIS for the overall leading method (inverse score with tuning), the popular mean benchmark and the leading model (Model 33) for forecasts at the individual state level.

**Fig 5.**
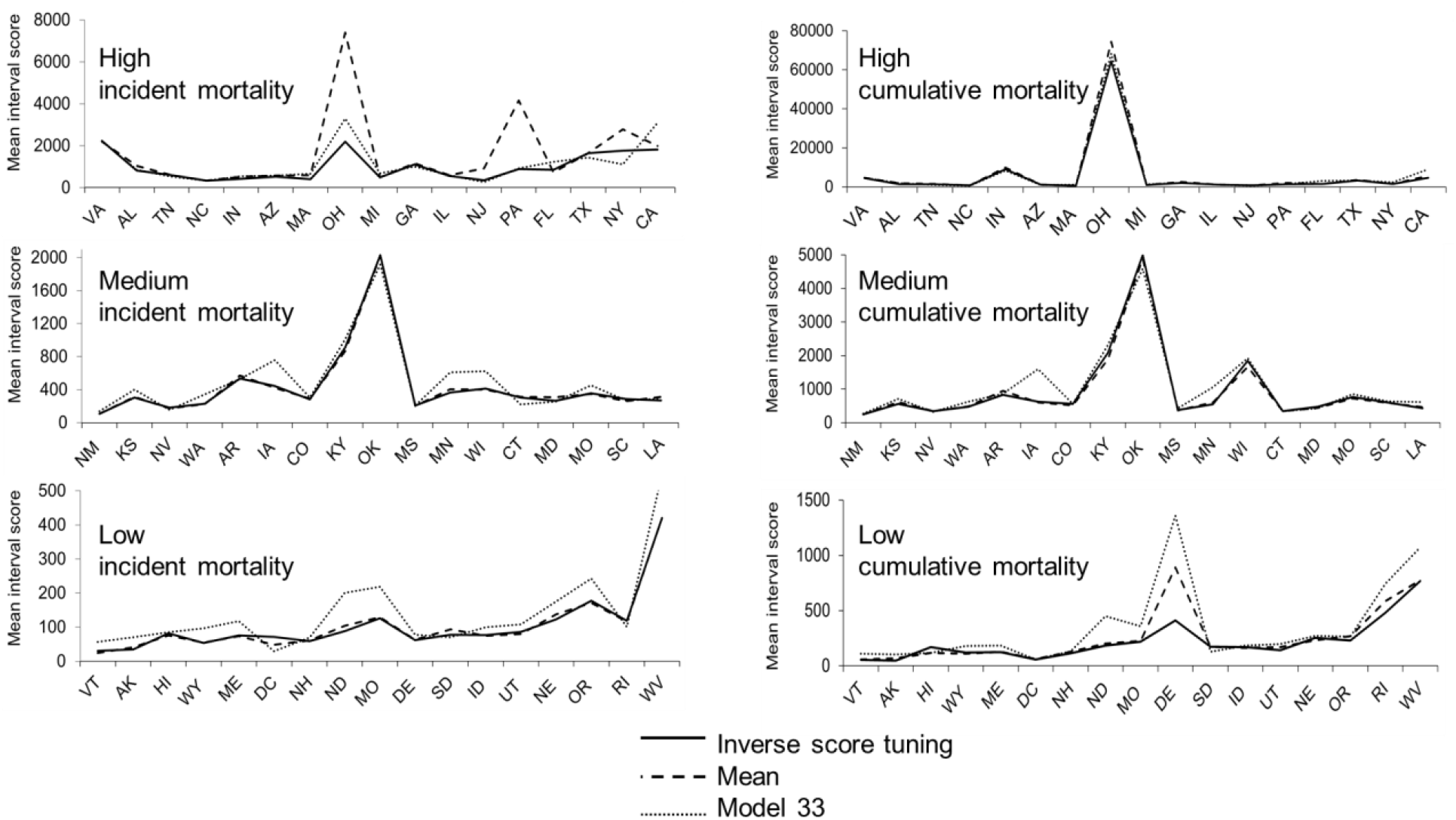
Mean interval scores for 95% interval forecasts of incident and cumulative COVID-19 mortalities in individual states in high, medium and low mortality groups, for three selected methods. The states are ordered on the x-axis by the number of cumulative COVID-19 deaths at the end of Week 73, from low to high. Lower scores reflect higher accuracy.

The adverse effects of reporting delays of death counts on performance is apparent for the high mortality state of Ohio (OH), the medium mortality state of Oklahoma (OK), and the low mortality states of Delaware (DE), West Virginia (WV) and Rhode Island (RI). For interval forecasts of both mortalities, the mean combination was competitive for most states. This is not reflected in Tables 1 to 2 due to adverse effects being greatest for the mean, particularly in the states of Ohio and Delaware. The impact of reporting delays on performance differed across mortalities. In particular, the inverse score with tuning method was less affected than the mean and Model 33 for interval forecasts of incident mortality in Ohio, whilst the effects were similar for interval forecasts of cumulative mortality (Fig 5). For the state of Delaware, the inverse score with tuning method was less affected for interval forecasts of cumulative mortality, but for interval forecasts of incident mortality, there was no effect on performance for all three methods (Fig 5). Similar adverse effects were reflected in the scores for the point forecasts (Fig 6).

**Fig 6.**
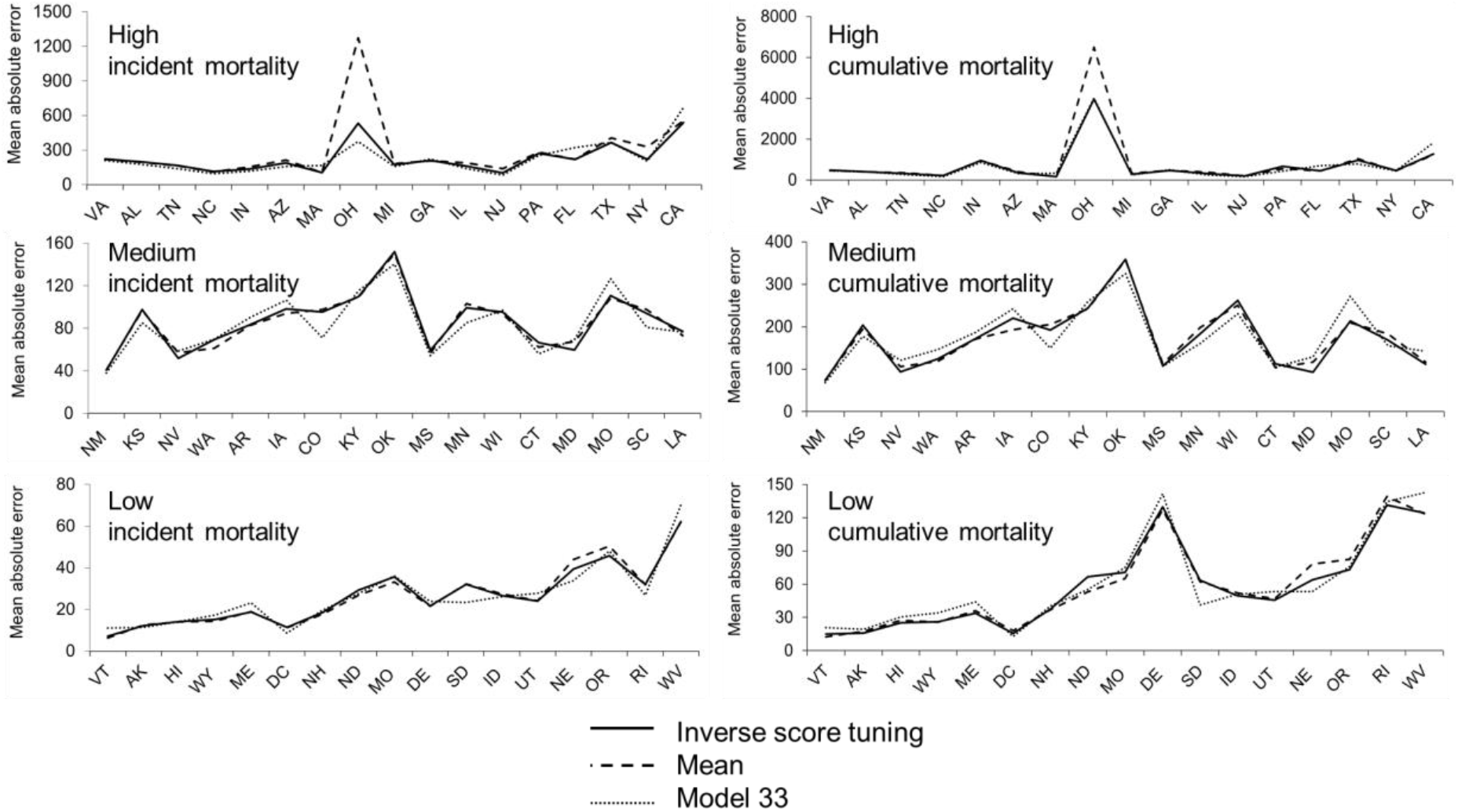
Mean absolute errors for median point forecasts of incident and cumulative COVID-19 mortalities in individual states in high, medium and low mortality groups, for three selected methods. The states are ordered on the x-axis by the number of cumulative COVID-19 deaths at the end of week 73 from low to high. Lower scores are better.

As a sensitivity analysis, we excluded forecasts for all six states that had notable effects of updates on death counts. There were improvements in the performance for all methods, and only slight changes in the ranking of methods. Regarding our significance testing results, there were no changes in our conclusions.

### Calibration of interval forecasting methods

There was a general tendency for underestimation in the widths of the 95% intervals (Table 5), which is quite common in studies of interval forecasting [43]. The inverse score methods were relatively well calibrated. Interior trimming was well calibrated, because it leads to wider than average intervals, which is useful when models underestimate the interval width. The envelope method also performed well because it also leads to wider than average intervals. However, this method is likely to deliver intervals that are too wide. Also, sizeable overestimation will lead to a result of 100% which is quite close to the ideal of 95%, while sizeable underestimation can lead to calibration far from 95%. This highlights a limitation of calibration for evaluating interval forecasts, and supports our greater focus on the interval score.

**Table 5.**
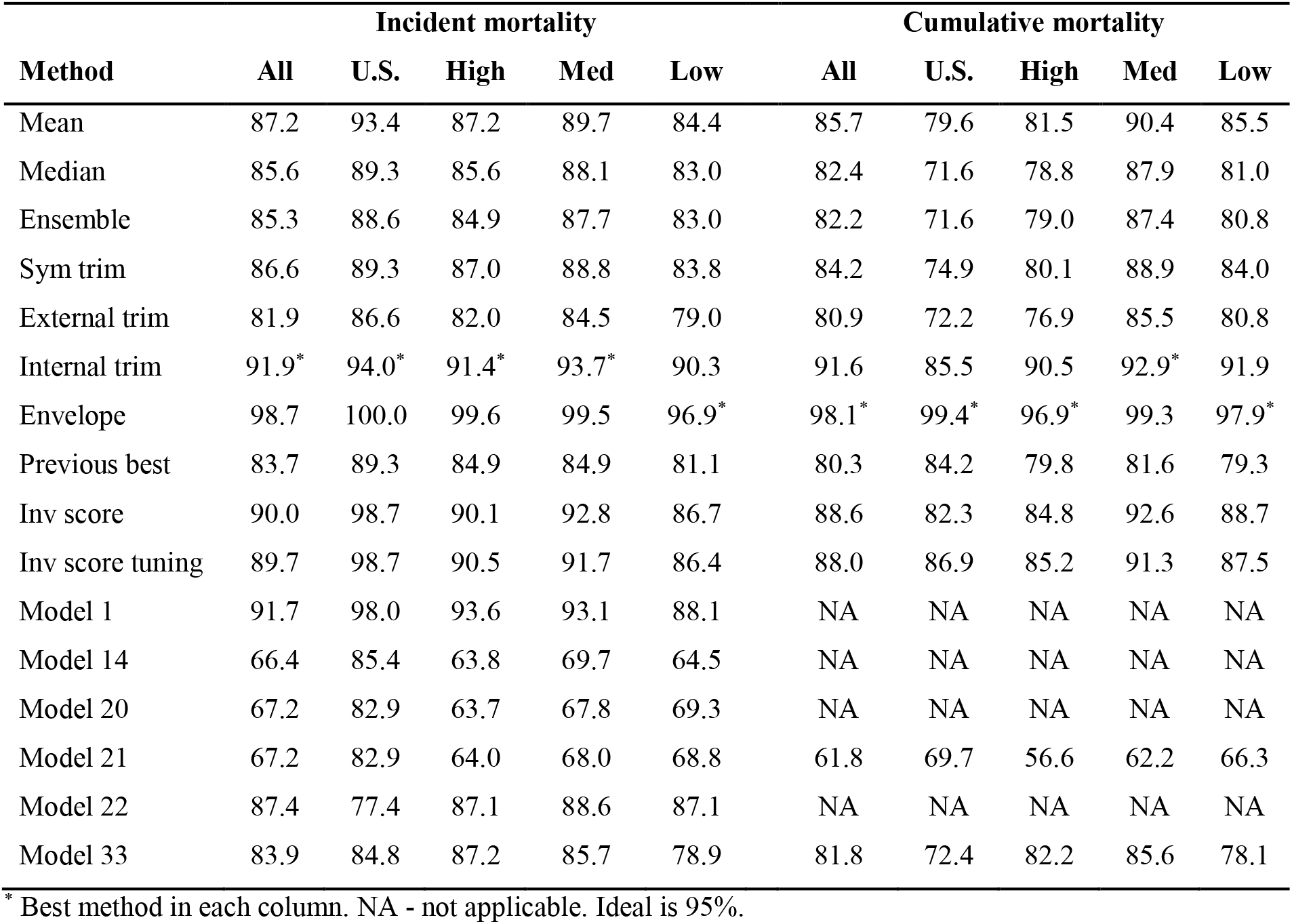
Average calibration of the 95% interval forecasts for incident and cumulative mortality in the 39 week out-of-sample period.

## Discussion

Our weighted combining methods performed well for both types of mortality, and for both interval and point forecasting. Inverse score with tuning was the most accurate method overall. The relative performance of the different combining methods, particularly the mean verses the median, depended on the mortality type, forecast type and timing. The leading individual model performed well, particularly for forecasts for the high mortality category, and this model was most accurate in point forecasting incident mortality. The adverse effects of reporting delays on performance were minor.

We developed the inverse score methods in our earlier study [32] and found them to be the most accurate combining methods overall. By formally testing the combining methods on an extended new dataset and a new dataset, drawing the same conclusion, our study provides further evidence of the superiority of these simple weighted methods. We also provide further insight into the relative performance of the simple benchmark methods. A recent study by Bracher et al [29] compared forecasts produced by the mean combination, median combination and a weighted combination for COVID-19 deaths in Germany and Poland. They found that combined methods did not perform better than individual models. However, this study was limited by an evaluation period of only ten weeks. It is also worth noting that the study used just thirteen individuals models in the combinations. In our previous work [32], we found accuracy improved notably as the number of individual models rose up to about twenty models.

We understand that this study is the first formal comparison of simple and more complex combining methods of probabilistic forecasts applied to infectious disease prediction and with a forecast evaluation period of at least 52 weeks. Another strength of our study is our source of data, which presented an opportunity to study the ‘wisdom of the crowd’, and provided the necessary conditions for the crowd being ‘wise’ [20] and without distortion, such as by social pressure [44] or restrictions against forecasting teams applying their own judgement [26]. These conditions include independent contributors, diversity of opinions, and a trustworthy central convener to collate the information provided [20]. Further strengths relating to the reliability of our findings arise from the high number of individual models included models in addition to the extended forecast evaluation period of our study. Our reported findings are limited to U.S. data and a particular set of models, and it is possible that different results may arise from other models, or for data from other locations, or other types of data, such as numbers infected. These are potential avenues for future research. Our ability to detect statistical differences was limited by the small sample sizes, with only 17 locations in each category, missing data and a relatively short out-of-sample period.

It is suggested that relying on modelling alone leads to “missteps and blind spots”, and that the best approach to support public policy decision making would involve a triangulation of insights from modelling with other information, such as analyses of previous outbreaks and discussions with frontline staff [45]. It is essential that modelling offers the most accurate forecasts. Probabilistic forecasts reflect the inherent uncertainties in prediction. Although individual models can sometimes be more accurate than combined methods, relying on forecasts from combined methods provides a more risk-averse approach, as the best individual model will not be clear until records of historical accuracy are available, and the best performing model will typically change over time. Furthermore, our finding that the performance of the average (mean combination) was substantially better than the average performance of the individual models suggests that, at the start of an epidemic, when it is not clear which model has the best performance, the statistical expectation is that the average method will score far better than a model chosen randomly, or chosen on the basis of no prior history, which was the case at the start of the COVID-19 pandemic. The performance of the mean and median combination were often reasonable, but there was no clear leading benchmark method, and when historical accuracy became available, the weighted methods were more accurate. Therefore, we only recommend considering simple methods in early stages of epidemics and we call for further formal investigation into the use and potential benefit of weighted methods in the prediction of diseases using probabilistic forecasts.

## Data Availability

This study is based on publically available data from the COVID-19 Forecast Hub.

https://github.com/reichlab/covid19-forecast-hub

## Acknowledgements

We are very grateful to Nia Roberts for clarifying our understanding of the license terms for the forecast data from the COVID-19 Forecast Hub.

## Supplementary information

**S1 File. Interval score combining methods**.

**S1 Table. Individual forecasting models**.

**S2 Table. For incident COVID-19 mortality, scores in the 39 week out-of-sample period for each horizon**.

**S3 Table. For cumulative COVID-19 mortality, scores in the 39 week out-of-sample period for each horizon**.

**S4 Table. For incident COVID-19 mortality, scores for each 13 week period**.

**S5 Table. For cumulative COVID-19 mortality, scores for each 13 week period**.

