## Supplemental File for "Forecasts of weekly incident and cumulative COVID-19 mortality in the United States: A comparison of combining methods"

**S1 File. Interval score combining methods**

*(a) Mean* combination. For each bound, we calculated the average of the forecasts of this bound.
*(b) Median* combination. For each bound, we calculated the median of the forecasts of this bound. This method is robust to outliers.
*(c) Symmetric trimming.* This method deals with outliers. For each bound, it involves trimming the $N$ lowest-valued and $N$ highest-valued forecasts, where $N$ is the largest integer less than or equal to the product of $\beta/2$ and the total number of forecasts. The median combination is an extreme form of symmetric trimming. The trimming parameter $\beta$ is optimised.
*(d) Exterior trimming*. This method addresses underconfidence, which is reflected by overly wide intervals. It involves removing the $N$ lowest-valued lower bound forecasts and the $N$ highest-valued upper bound forecasts, where $N$ is the largest integer less than or equal to the product of $\beta$ and the number of forecasts. When trimming resulted in a lower bound being above the upper bound, we replaced the two bounds by their average. The trimming parameter $\beta$ is optimised.
(e) *Interior trimming*. This method addresses overconfidence, which is reflected by overly narrow intervals. It involves removing the $N$ highest-valued lower bound forecasts and the $N$ lowest-valued upper bound forecasts, where $N$ is defined as for exterior trimming. The trimming parameter $\beta$ is optimised.
(f) *Envelope method*. The interval is constructed using the lowest-valued lower bound forecast and highest-valued upper bound forecast. This method is an extreme form of interior trimming.
(g) *Ensemble* - COVID-19 Hub's ensemble forecast. This was initially the mean combination of the forecasts that they considered to be eligible, but in late July, 12 weeks into our 52-week analysis period, it became the median combination of these forecasts. The use of eligibility screening implies that the ensemble is constructed with the benefit of a degree of subjective trimming.
(h) P*revious best.* This is the model with the best historical accuracy at the current forecast origin. For this method, the interval forecast was obtained from the model for which the MIS was the lowest when computed using the weeks up to and including the current forecast origin (i.e., the in-sample period).
(i) *Inverse interval score* method. This is a weighted method using historical accuracy, with weights inversely proportional to the mean interval score (MIS).
(j) *Inverse interval score with tuning*. This is a second weighted method, with weights inversely proportional to the MIS and a tuning parameter, $\lambda > 0$, incorporated to control the influence of the score on the combining weights. The following expression gives the weight on forecasting model $i$ at forecast origin $t$:

$$w_{it} = \frac{\left({}^{1}/_{MIS_{i,t}}\right)^{\lambda}}{\sum_{j=1}^{J}\left({}^{1}/_{MIS_{i.t}}\right)^{\lambda}}$$

where $MIS_{i,t}$ is the historical MIS computed at forecast origin $t$ from model $i$, and $J$ is the number of forecasting models included in the combination. If $\lambda$ is close to zero, the combination reduces to the mean combination, whereas a large value for $\lambda$ leads to the selection of the model with best historical accuracy. The parameter $\lambda$ was optimised using the same expanding in-sample periods, as for the trimming combining methods. Due to the extent of missing forecasts, we pragmatically computed $MIS_{i,t}$ using all available past forecasts, rather than limit the computation to periods for which forecasts from all models were available. For the models for which forecasts were not available for at least 5 past periods, we set $MIS_{i,t}$ to be equal to the mean of $MIS_{i,t}$ for all other models. An alternative approach, which we employed in our earlier study, is to omit from the combination any model for which there is only a very short or non-existent history of accuracy available. The disadvantage of this is that it omits potentially useful forecast information, and this was shown by our empirical forecasting results.
