## Supplemental Table 1 for "Forecasts of weekly incident and cumulative COVID-19 mortality in the United States: A comparison of combining methods"

**S1 Table. Individual forecasting models**

| Contributors | Short model name | Model description* | Access and licencing information Citations |
|---|---|---|---|
| Wattanachit N, Ray EL, Reich N | COVID hub-ensemble | An ensemble, or model average, of submitted forecasts to the COVID-19 Forecast Hub. | https://github.com/reichlab/covid19-forecast-hub/tree/master/data-processed/COVIDhub-ensemble https://www.medrxiv.org/content/10.1101/2020.08.19.20177493v1 |
| *COMPARTMENTAL* | | | |
| Tomar V, Jain C | Auquan-SEIR† | Modified SEIR model with compartments for reported and unreported infections. Non-linear mixed effects curve-fitting. | https://github.com/reichlab/covid19-forecast-hub/tree/master/data-processed/Auquan-SEIR |
| Panano B. | BPangano-RtDriven | Projects infections and deaths for 223 locations using an SIR model. | https://github.com/reichlab/covid19-forecast-hub/tree/master/data-processed/BPagano-RtDriven<br><br>https://bobpagano.com/covid-19-modeling/ |
| Carlson E, Henderson M, Kelly C, Kofman I, Zhang X | CovidActNow-SEIR_CAN | SEIR model forecasts of cumulative deaths, incident deaths, incident hospitalizations by fitting predicted cases, deaths, and hospitalizations to the observations. | https://github.com/reichlab/covid19-forecast-hub/tree/master/data-processed/CovidActNow-SEIR_CAN |
| Li ML, Bouardi HT, Lami OS, Trikalinos TA, Trichakis NK, Bertsimas D | CovidAnalytics-DELPHI | SEIR model augmented with underdetection and interventions. Projections account for reopening and assume interventions would be re-enacted if cases continue to climb. | https://github.com/reichlab/covid19-forecast-hub/tree/master/data-processed/CovidAnalytics-DELPHI https://www.covidanalytics.io/DELPHI_documentation_pdf |
| Chhatwal J, Ayer T, Linas B, Dalgic O, Mueller P, Adee M, Ladd MA, Xiao J | Covid19Sim-Simulator | An interactive tool that uses a validated SEIR compartment model. | https://github.com/reichlab/covid19-forecast-hub/tree/master/data-processed/Covid19Sim-Simulator |
| Pei S, Yamana T, Kandula S, Yang W, Galanti M, Shaman J | CU-select | Metapopulation county-level SEIR model for projecting future COVID-19 incidence and deaths. This forecast is the scenario we believe to be most plausible given the current setting. | https://github.com/reichlab/covid19-forecast-hub/tree/master/data-processed/CU-select https://doi.org/10.1101/2020.03.21.20040303 https://www.medrxiv.org/content/10.1101/2020.05.04.20090670v2 |
| Pei S, Yamana T, Kandula S, Yang W, Galanti M, Shaman J | CU-nochange | This metapopulation county-level SEIR model assumes that current contact rates will | https://github.com/reichlab/covid19-forecast-hub/tree/master/data-processed/CU-nochange |

| | | | |
|---|---|---|---|
| | | remain unchanged in the future. | https://doi.org/10.1101/2020.03.21.20040303 |
| Max A, Epshteyn A, Kang B, Li C-L, Sava D, Parish D, Miller D, Kanal E, Liu H, Nakhost H, Jones I, Lai J, Repenning J, Yoon J, Ramasamy K, Zhang L, Le L, Nikoltchev M, Siegler M, Dusenberry M, Yoder N, Rozenfeld O, Rangaswamy P, Sinha R, Xie R, Arik S, Singh S, Tsai T, Pfister T, Menon V, Karande V, Y, Li Y | Google-Harvard-CPF | Our model improves upon standard compartmental models by using temporally and spatially rich data, and integrating covariate encodings into compartment transitions via end-to-end learning. | https://github.com/reichlab/covid19-forecast-hub/tree/master/data-processed/Google_Harvard-CPF https://arxiv.org/abs/2008.00646 |
| Lemaitre JC, Bi Q , Hulse JD, Grabowski MK, Grantz KH, Kaminsky J, Lauer SA, Lee EC, Meredith HR, Perez-Saez J, Truelove SA, Keegan LT, Kaminsky K, Shah S, Wills J, Aquilanti P-Y, Raman K, Subramaniyan A, Thursam G, Tran A. | JHU_IDD-CovidSP | County-level metapopulation model with commuting and stochastic SEIR disease dynamics with social-distancing indicators. | https://github.com/reichlab/covid19-forecast-hub/tree/master/data-processed/JHU_IDD-CovidSP https://doi.org/10.1038/s41598-021-86811-0 |
| Kinsey M, Tallaksen K, Obrecht RF, Asher L, Costello C, Kelbaugh M, Wilson S | JHUAPL_Bucky | Metapopulation model using public mobility data. Local parameters (case reporting rates, doubling times, etc) are estimated using data from CSSE and CDC scenario 5. Primary output is case incidence. | https://github.com/reichlab/covid19-forecast-hub/tree/master/data-processed/JHUAPL-Bucky |
| Baek J, Farias V, Georgescu A, Levi R, Sinha D, Wilde J, Zheng A | MITCovAlliance-SIR | SIR model trained on public heath regions. SIR parameters are functions of static demographic and time-varying mobility features. Two-stage approach that first learns magnitude of peak infections. | https://github.com/reichlab/covid19-forecast-hub/tree/master/data-processed/MITCovAlliance-SIR https://arxiv.org/abs/2006.06373 |
| Vespignani A, Chinazzi M, Davis JT, Mu K, Pastore y Piontti A, Samay N, Xiong X, Halloran ME, Longini IM, Dean NE, Viboud C, Sun K, Litvinova M, | MOBS-GLEAM_COVID | Metapopulation, age structured SLIR model. Superimposed on the worldwide population and mobility layers is an agent-based epidemic model that defines the infection and population dynamics. Makes predictions about the future | https://github.com/reichlab/covid19-forecast-hub/tree/master/data-processed/MOBS-GLEAM_COVID https://uploads-ssl.webflow.com/58e6558acc00ee8e4536c1f5/5e8bab44f5baae4c1c2a75d2_GLEAM_web.pdf |

| | | | |
|---|---|---|---|
| Gioannini C, Rossi L, Ajelli M | | that are dependent on the assumption that current interventions continue. | |
| Gao Z, Li C, Zheng S, Bian J, Xie X, LiuT-Y | MSRA-DeepST | A deep spatio-temporal network with knowledge based SEIR as a regularier under the assumption of spatio-temporal process in pandemic of different regions. | https://github.com/reichlab/covid19-forecast-hub/tree/master/data-processed/MSRA-DeepST |
| Espana G, Oidtman R, Cavany S, Costello A, Wieler A, Lerch A, Barbera C, Poterek M, Tran Q, Moore S, Perkins A | NotreDame-Mobility | Ensemble of nine models that are identical except that they are driven by different mobility indices from Apple and Google. The model underlying each is a deterministic, SEIR-like model. | https://github.com/reichlab/covid19-forecast-hub/tree/master/data-processed/NotreDame-mobility |
| Koyluoglu U, Milliken J | OliverWyman-Navigator | Forecasts and scenario analysis for Detected and Undetected cases and death counts following a compartmental formulation with non-stationary transition rates. | https://github.com/reichlab/covid19-forecast-hub/tree/master/data-processed/OliverWyman-Navigator |
| Turtle J, Ben-Nun M, Riley P | PSI-DRAFT | A stochastic/deterministic, single-population SEIRX model that stratifies by both age distribution and disease severity and includes generic intervention fitting. | https://github.com/reichlab/covid19-forecast-hub/tree/master/data-processed/PSI-DRAFT |
| Shi Y, Shah T, Ban X | RPI-UW-Mob_Collision | A mobility-informed simplified SIR model motivated by collision theory. | https://github.com/reichlab/covid19-forecast-hub/tree/master/data-processed/RPI-UW-Mob-Collision https://www.medrxiv.org/content/10.1101/2020.07.25.20162016v1 |
| Snyder TL, Wilson DD | SWC-TerminusCM | Mechanistic compartmental model using disease parameter estimates from literature. It uses Bayesian inference to predict the most likely model parameters. | https://github.com/reichlab/covid19-forecast-hub/tree/master/data-processed/SWC-TerminusCM |
| Cobey S, Arevalo P, Baskerville E, Carran S, Gostic K, McGough L, Ranjeva S, Wen F | UChicago-COVIDIL | Compartmental, age-structured SEIR model that infers past SARS-CoV-2 transmission rates and forecasts mortality under current and hypothetical public health interventions. | https://github.com/reichlab/covid19-forecast-hub/tree/master/data-processed/UChicago-CovidIL |
| Gu Q, Xu P, Chen J, Wang L, Zou D, Zhang W | UCLA-SuEIR | Variant of the SEIR model considering both untested and unreported cases. The model considers reopening and assumes susceptible | https://github.com/reichlab/covid19-forecast-hub/tree/master/data-processed/UCLA-SuEIR |

| | | | |
|---|---|---|---|
| | | population will increase after the reopen. | https://www.medrxiv.org/content/10.1101/2020.05.24.20111989v1 |
| Chen YQ, Zhao Y, Guo L | UCM-MESALab-FoGSEIR | FoGSEIR model is a modification of integer order SEIR model considering fractional integrals. The model considers the age structure and reopening intervention to minimize infections and deaths. | https://github.com/reichlab/covid19-forecast-hub/tree/master/data-processed/UCM_MESALab-FoGSEIR |
| Sheldon D, Gibson G, Reich N | UMass-MechBayes | Bayesian compartmental model with observations on cumulative case counts and cumulative deaths. Model is fit independently to each state. Model includes observation noise and a case detection rate. | https://github.com/reichlab/covid19-forecast-hub/tree/master/data-processed/UMass-MechBayes |
| Mayo ML, Rowland MA, Parno MD, Detwiller ID, Farthing MW, England WP George GE | USACE-ERDC_SEIR | The ERDC SEIR model makes predictions of several variables (e.g., reported new/cumulative cases per day, etc.). Model parameters are estimated using historical data using Bayesian inference. | https://github.com/reichlab/covid19-forecast-hub/tree/master/data-processed/USACE-ERDC_SEIR |
| Jain S, Tiwari A, Deva A, Kulkarni M, Shingi S, Bannur N, White J, Merugu S, Raval A | Wadhwani_AI-BayesOpt | A novel model-agnostic Bayesian optimization ("BayesOpt") approach for learning the parameters of our SEIR model from observed data. | https://github.com/reichlab/covid19-forecast-hub/tree/master/data-processed/Wadhwani_AI-BayesOpt |
| Gu Y | YYG-ParamSearch | Based on the SEIR model with hyperparameter optimization to make daily projections regarding COVID-19 infections and deaths in 50 US states. The model accounts for state reopenings and its effects on infections and deaths. | https://github.com/reichlab/covid19-forecast-hub/tree/master/data-processed/YYG-ParamSearch https://covid19-projections.com/about/ |
| *NON-COMPARTMENTAL* | | | |
| O'Dea E | CEID-Walk | A random walk model with drift. A least squares line is fitted to the tail observations of a target time series to estimate the drift and step variance of a random walk model. | https://github.com/reichlab/covid19-forecast-hub/blob/master/data-processed/CEID-Walk/metadata-CEID-Walk.txt |
| Green A, Hu A, Jahja M, Ventura V, Wasserman L, Tibshirani Rob, Shankar V, Bien J, Brooks L, | CMU-Timeseries § | A basic AR-type time series model fit using case counts and deaths as features. | https://github.com/reichlab/covid19-forecast-hub/tree/master/data-processed/CMU-TimeSeries |

| | | | |
|---|---|---|---|
| Narasimhan B, Rajanala S, Rumack A, Simon N, Sharpnack J, McDonald D(University of British Columbia), Ryan Tibshirani (Senior author, and the Delphi COVID-19 Response Team | | | |
| Wang Y, Zeng D, Wang Q, Xie S | Columbia_UNC-SurvCon | Survival-convolution model with piece-wise transmission rates that incorporates latent incubation period and provides time-varying effective reproductive number. | https://github.com/reichlab/covid19-forecast-hub/tree/master/data-processed/Columbia_UNC-SurvCon https://www.frontiersin.org/article/10.3389/fpubh.2020.00325 |
| Ray EL, Tibshirani R | COVIDhub-baseline | Baseline prediction model. | https://github.com/reichlab/covid19-forecast-hub/tree/master/data-processed/COVIDhub-baseline |
| Kalantari R, Zhou M. | DDS-NBDS | Jointly modeling daily deaths and cases using a negative binomial distribution based nonparametric Bayesian generalized linear dynamical system. | https://github.com/reichlab/covid19-forecast-hub/tree/master/data-processed/DDS-NBDS https://dds-covid19.github.io/ |
| Sherratt K, Bosse N, Abbott S, Hellewell J, Meakin S, Munday J, Funk S | epiforecasts-ensemble1 | A deaths forecast using the renewal equation and time-series forecasts of the time-varying reproduction number. | https://github.com/reichlab/covid19-forecast-hub/tree/master/data-processed/epiforecasts-ensemble1 https://doi.org/10.12688/wellcomeopenres.16006.1 |
| Keskinocak P, Aglar BEO, Baxter A, Asplund J, Serban N | GT_CHHS-COVID19 | Agent-based simulation model to project COVID19 infection spread. | https://github.com/reichlab/covid19-forecast-hub/tree/master/data-processed/GT_CHHS-COVID19 |
| Prakash BA, Rodriguez A, Cui J, Tabassum A, Adhikari B, Sun J, Xiao D, Qiang C | GT-DeepCOVID | Data-driven approach based on deep learning for forecasting mortality and hospitalizations using syndromic, clinical, demographic, mobility and point-of-care data. | https://github.com/reichlab/covid19-forecast-hub/tree/master/data-processed/GT-DeepCOVID |
| Murry C and the IHME-CurveFitTeam | IHME-CurveFit | Non-linear mixed effects curve-fitting. This model makes predictions about the future that are dependent on the assumption that current interventions continue. | https://github.com/reichlab/covid19-forecast-hub/tree/master/data-processed/IHME-CurveFit https://www.medrxiv.org/content/10.1101/2020.03.27.20043752v1 |

| | | | |
|---|---|---|---|
| Wang L, Wang G, Gao L, Li X, Yu S, Kim M, Wang Y, Gu Z. | IowaStateLW-STEM | A nonparametric space-time disease transmission model. The projections assume that the data used is reliable, the future will continue to follow the current pattern, and current interventions will remain the same till the end of forecasting period. | https://github.com/reichlab/covid19-forecast-hub/tree/master/data-processed/IowaStateLW-STEM https://arxiv.org/abs/2004.14103 |
| Chiang W-H, Mohler G | IUPUI-HkPrMobiDyR | Hawkes processes with Dynamic reproduce number. | https://github.com/reichlab/covid19-forecast-hub/tree/master/data-processed/IUPUI-HkPrMobiDyR<br><br>https://doi.org/10.1101/2020.06.06.20124149 |
| Marshall M, Gardner L, Drew C, Burman E, Nixon K | JHU_CSSE-DECOM | County-level, empirical machine learning model driven by epidemiological, mobility, demographic, and behavioral data. | https://github.com/reichlab/covid19-forecast-hub/tree/master/data-processed/JHU_CSSE-DECOM |
| Karlem D | Karlen-pypm | Discrete-time difference equations with long periods of constant transmission rate | https://github.com/reichlab/covid19-forecast-hub/tree/master/data-processed/Karlen-pypm<br><br>https://arxiv.org/abs/2007.07156 |
| Osthus D, Del Valle S, Manore C, Weaver B, Castro L, Shelley S, Smith M, Spencer J, Fairchild G, Travis Pitts T, Gerts D, Dauelsberg L, Daughton A, Gorris M, Hornbein B, Israel D, Parikh N, Shutt D, Ziemann A | LANL-GrowthRate | Statistical dynamical growth model accounting for population susceptibility. Makes predictions about the future, unconditional on particular intervention strategies. | https://github.com/reichlab/covid19-forecast-hub/tree/master/data-processed/LANL-GrowthRate |
| Gao Z, Li C, Cao W, Zheng S, Bian J, Xie X, Liu TY, Zhang S, Ferres JL | Microsoft-DeepSTIA† | A deep spatio-temporal network with intervention and hospital gate under the assumption of spatio-temporal process in pandemic of different regions. | https://github.com/reichlab/covid19-forecast-hub/tree/master/data-processed/Microsoft-DeepSTIA |
| Espana G, Oidtman R, Cavany S, Costello A, Wieler A, Lerch A, Barbera C, Poterek M, Tran Q, Moore S, Perkins A | NotreDame-FRED | Agent-based model developed for influenza with parameters modified to represent the natural history of COVID-19 | https://github.com/reichlab/covid19-forecast-hub/tree/master/data-processed/NotreDame-FRED |
| Walraven R | RobertWalraven-ESG | Multiple skewed gaussian distribution peaks fitted to raw data. | https://github.com/reichlab/covid19-forecast-hub/tree/master/data-processed/RobertWalraven-ESG |

| | | | |
|---|---|---|---|
| Nagraj VP, Turner SD, Hulme-Lowe C | SigSci_TS | Time series forecasting using ARIMA for case forecasts and lagged cases for death forecasts. | https://github.com/reichlab/covid19-forecast-hub/tree/master/data-processed/SigSci-TS |
| McConnell S, Donaldson B | SteveMcConnell_ COVIDComplete | A near-term fatality prediction model that calculates and uses fatality trends at the national and state level, trends in positive virus tests and total virus tests, and age-related demographics for state forecasts. Model forecasts are based on predicting near-term deaths from recent positive virus tests. | https://github.com/reichlab/covid19-forecast-hub/tree/master/data-processed/SteveMcConnell-CovidComplete https://stevemcconnell.com/covid |
| Bieggel H, Lega J | UA-EpiCovDA | SIR mechanistic model with data assimilation. EpiCovDA is an extension of the EpiGro model. Model parameters are fit to Covid-19 data using a variational data assimilation method. A prior distribution of the parameters is estimated by fitting an SIR Incidence-Cumulative Cases curve to data from states that had at least 1000 cases by 04/01/2020. | https://github.com/reichlab/covid19-forecast-hub/tree/master/data-processed/UA-EpiCovDA |
| Jin X, Wang Y-X, Yan X | UCSB-ACTS | This data-driven machine learning model makes predictions by referring to other regions with similar growth patterns and assuming the similar development will take place in the current region. | https://github.com/reichlab/covid19-forecast-hub/tree/master/data-processed/UCSB-ACTS |
| Wu D, Gao L, M Yian, Yu R, Vespignani A, Chinazzi M, Davis JT, Mu K, Pastore y Piontti A, Xiong X | UCSD-NEU_DeepGLEAM | Combines the signal of a discrete stochastic epidemic computational model GLEAM with a deep learning spatiotemporal forecasting framework to further improve predictions.' | https://github.com/reichlab/covid19-forecast-hub/tree/master/data-processed/UCSD_NEU-DeepGLEAM |
| Corsetti S, Schwarz T | UMich-RidgeTfReg | Nation-level model of confirmed cases and deaths based on ridge regression. No assumptions made about social distancing. | https://github.com/reichlab/covid19-forecast-hub/tree/master/data-processed/UMich-RidgeTfReg |
| Zhang-James Y, Hess J, Chen S, Wang D, Morley CP, Faraone SV. | UpstateSU_GRU § | County-level forecast using recurrent neural network seq2seq model with the Gated recurrent units (GRU) | https://github.com/reichlab/covid19-forecast-hub/tree/master/data-processed/UpstateSU-GRU |
| Srivastava A, Prasanna VK, Xu FT | USC-SI_kJalpha § | A heterogeneous infection rate model with human | https://github.com/reichlab/covid19-forecast- |

| | | | |
|---|---|---|---|
|  |  | mobility for epidemic modeling. Our model adapts to changing trends and provide predictions of confirmed cases and deaths. | hub/tree/master/data-processed/USC-SI_kJalpha<br><br>https://arxiv.org/abs/2007.05180 |
| Srivastava A, Prasanna VK, Xu FT | USC-SI_kJalpha_RF | A heterogeneous infection rate model with human mobility for epidemic modeling. Our model adapts to changing trends and provide predictions of confirmed cases and deaths. We build a random forest, based on the output of USC_SIkJalpha model along with the data on the cumulative case/death, weekly increase, and previous increase. We then sample trees to generate quantile forecasts | https://github.com/reichlab/covid19-forecast-hub/tree/master/data-processed/USC-SI_kJalpha_RF<br><br>https://arxiv.org/abs/2007.05180 |
| Woody S, et al. at the University of Texas | UT-Mobility | This model makes predictions assuming that social distancing patterns, as measured by anonymized mobile-phone GPS traces, remain constant in the future. Only models *first-wave deaths*. | https://github.com/reichlab/covid19-forecast-hub/tree/master/data-processed/UT-Mobility |
| Mehrotra P, Ivan JI, and the Walmart Labs COVID-19 Team | WalmartLabsML_LogForecasting† | A logistic growth prophet forecasting model fit using case counts and deaths as features.The Model is built by Prophet model with logistic growths to forecast the US cumulative deaths. By sampling from uniform distribution to get the quantiles. | https://github.com/reichlab/covid19-forecast-hub/tree/master/data-processed/WalmartLabsML-LogForecasting |

* Based on information recorded on the COVID19 Hub with citations as recorded on 18/5/21; † Only provided forecasts of numbers of cumulative COVID-19 deaths; § Only provided forecasts of numbers of incident COVID-19 deaths.
