## Supplemental Table 2 for "Forecasts of weekly incident and cumulative COVID-19 mortality in the United States: A comparison of combining methods"

**S2 Table 1. For incident COVID-19 mortality, scores in the 39 week out-of-sample period for each horizon**

| Method | Mean interval score | | | | | Mean absolute error | | | | |
|---|---|---|---|---|---|---|---|---|---|---|
| | All | U.S. | High | Med | Low | All | U.S. | High | Med | Low |
| *1 week ahead* | | | | | | | | | | |
| Mean | 687 | 6877 | 1224 | 386 | 88 | 68 | 1202 | 92 | 34 | 11 |
| Median | 618 | 8679 | 914 | 370 | 95 | 64 | 1152 | 86 | 33 | 11 |
| Ensemble | 621 | 8541 | 927 | 374 | 95 | 65 | 1181 | 85 | 33 | 11 |
| Sym trim | 638 | 7222 | 1075 | $364^{\dagger}$ | $88^{\S}$ | $65^{\dagger}$ | 1152 | 87 | $32^{*\dagger}$ | 11 |
| Interior trim | 647 | 6999 | 1125 | $361^{*}$ | $81^{*\dagger\S}$ | 68 | 1223 | 91 | 33 | 11 |
| Inv score | $594^{\dagger}$ | 6506 | $988^{\dagger}$ | $364^{\dagger}$ | $84^{\dagger\S}$ | $66^{\dagger}$ | 1159 | 89 | 33 | 11 |
| Inv score tuning | $533^{\dagger}$ | 6245 | 807 | 370 | $87^{\S}$ | $62^{*\dagger}$ | $1032^{*}$ | $85^{\dagger}$ | $34^{\dagger}$ | $11^{\dagger}$ |
| Model 21 | $527^{*\dagger}$ | $6005^{*}$ | $770^{*\S}$ | 394 | 94 | $62^{*}$ | 1039 | $84^{*}$ | 33 | $10^{*}$ |
| Model 33 | 710 | 8352 | 1121 | 449 | 112 | 66 | 1081 | 93 | 34 | $10^{*}$ |
| *2 weeks ahead* | | | | | | | | | | |
| Mean | 892 | 9345 | 1644 | 436 | 97 | 99 | 1486 | 162 | 40 | 13 |
| Median | 705 | 9795 | 1072 | $407^{*}$ | 100 | 76 | 1385 | $102^{*}$ | $38^{*\dagger}$ | $12^{*\dagger}$ |
| Ensemble | 713 | 9739 | 1087 | 418 | 101 | 77 | 1426 | $102^{*}$ | $38^{*\dagger}$ | $12^{*}$ |
| Sym trim | 841 | 9761 | 1477 | 424 | 98 | $94^{\dagger}$ | 1390 | 156 | $38^{*\dagger}$ | $12^{*\dagger}$ |
| Interior trim | 913 | 9322 | 1743 | 412 | $91^{*}$ | 99 | 1514 | 161 | 39 | 13 |
| Inv score | $742^{\dagger}$ | 8069 | $1285^{\dagger}$ | 418 | $91^{*\dagger}$ | 96 | $1385^{\dagger}$ | 161 | 39 | $12^{*}$ |
| Inv score tuning | $635^{*\dagger}$ | $7693^{*}$ | 958 | 434 | 96 | 75 | $1162^{\dagger}$ | 108 | 40 | $12^{*}$ |
| Model 21 | 645 | 8660 | $888^{*}$ | 473 | 102 | 74 | 1237 | 103 | 40 | $12^{*}$ |
| Model 33 | 720 | 9218 | 1088 | 454 | 120 | ***73*** | $1155^{*}$ | 105 | $38^{*}$ | $12^{*}$ |
| *3 weeks ahead* | | | | | | | | | | |
| Mean | 979 | 9660 | 1863 | 459 | 105 | 107 | 1809 | 163 | 46 | 14 |
| Median | 799 | 12427 | 1160 | 446 | 107 | 92 | 1726 | 124 | $43^{*\dagger}$ | 14 |
| Ensemble | 813 | 11915 | 1217 | 459 | 108 | 92 | 1708 | 124 | 44 | 14 |
| Sym trim | 953 | 11156 | 1694 | 456 | 110 | 103 | 1722 | 157 | 44 | 14 |
| Interior trim | 942 | 9921 | 1751 | 447 | $100^{*}$ | 107 | 1825 | 162 | 45 | 14 |
| Inv score | $815^{\dagger}$ | 8952 | $1432^{\dagger}$ | $436^{*}$ | $100^{*\dagger}$ | $103^{\dagger}$ | $1666^{\dagger}$ | 159 | 45 | $14^{\dagger}$ |
| Inv score tuning | $686^{*}$ | 8912 | 1018 | 452 | 105 | 88 | 1451 | 124 | 46 | 14 |
| Model 21 | 806 | 13889 | $988^{*}$ | 548 | 113 | 90 | 1632 | $119^{*}$ | 46 | $13^{*}$ |
| Model 33 | 698 | $6844^{*}$ | 1097 | 495 | 140 | $80^{*}$ | $1191^{*}$ | $119^{*}$ | $43^{*}$ | 14 |
| *4 weeks ahead* | | | | | | | | | | |
| Mean | 1044 | 13315 | 1792 | 491 | 126 | 121 | 2347 | 165 | 52 | 16 |
| Median | 989 | 14055 | 1543 | 518 | 138 | 112 | 2263 | 143 | $49^{*}$ | 15 |
| Ensemble | 1029 | 14509 | 1611 | 539 | 143 | 111 | 2204 | 145 | 50 | 15 |
| Sym trim | 1077 | 13641 | 1814 | 542 | 138 | 118 | 2263 | 163 | 51 | 15 |
| Interior trim | 1076 | 13634 | 1879 | 495 | $116^{*\dagger}$ | 121 | 2353 | 165 | 52 | 16 |
| Inv score | $929^{\dagger}$ | 12993 | $1478^{\dagger}$ | $481^{*}$ | 117 | $116^{\dagger}$ | $2130^{\dagger}$ | 162 | 51 | 16 |
| Inv score tuning | $854^{*\S}$ | 12906 | 1216 | 508 | 128 | $104^{\dagger}$ | 1777 | 145 | 53 | 15 |
| Model 21 | 939 | 16141 | $1145^{*}$ | 632 | 146 | 106 | 1931 | 141 | 55 | $14^{*}$ |
| Model 33 | 863 | $10312^{*}$ | 1249 | 598 | 185 | $99^{*}$ | $1680^{*}$ | $137^{*}$ | 51 | 17 |

$^{*}$For each horizon, best method for each column; $^{\dagger}$score is significantly lower than the mean combination; $^{\S}$ score is significantly lower than the median combination. Lower values are better.
