## Supplemental Table 3 for "Forecasts of weekly incident and cumulative COVID-19 mortality in the United States: A comparison of combining methods"

**S3 Table. For cumulative COVID-19 mortality, scores in the 39 week out-of-sample period for each horizon**

| | Mean interval score | | | | | Mean absolute error | | | | |
|---|---|---|---|---|---|---|---|---|---|---|
| **Method** | **All** | **U.S.** | **High** | **Med** | **Low** | **All** | **U.S.** | **High** | **Med** | **Low** |
| *1 week ahead* | | | | | | | | | | |
| Mean | 3319 | 62108 | 5666 | 585 | 249 | 160 | 3541 | 223 | 40 | 17 |
| Median | 3123 | 57217 | 5473 | 451$^{*\dagger}$ | 263 | 144$^{\dagger}$ | 3036 | 209$^{*\dagger}$ | 36$^{*\dagger}$ | 16$^{*\dagger}$ |
| Ensemble | 3077 | 54979 | 5461 | 453$^{\dagger}$ | 265 | 143$^{\dagger\S}$ | 2979 | 209$^{*\dagger}$ | 36$^{*\dagger}$ | 16$^{*\dagger}$ |
| Sym trim | 3277 | 63747 | 5560 | 483$^{\dagger}$ | 231$^{\S}$ | 148 | 3190 | 213 | 37$^{*\dagger}$ | 16$^{*\dagger}$ |
| Interior trim | 2204$^{*\dagger\S}$ | 33641$^{\dagger\S}$ | 4136$^{*\dagger\S}$ | 491$^{\dagger}$ | 136$^{*\dagger\S}$ | 156 | 3402 | 220 | 39 | 17$^{\dagger}$ |
| Previous best | 2559$^{\dagger\S}$ | 31436$^{*\S}$ | 5282 | 532 | 165$^{\dagger\S}$ | 147 | 2767$^{*\dagger}$ | 228 | 44 | 17 |
| Inv score | 3195 | 55789$^{\dagger}$ | 5681 | 612 | 199$^{\dagger\S}$ | 159 | 3548 | 222 | 40 | 16$^{*\dagger}$ |
| Inv score tuning | 2736$^{\dagger\S}$ | 39628$^{\S}$ | 5290$^{\dagger}$ | 574 | 175$^{\dagger\S}$ | 158 | 3520 | 219 | 40 | 16$^{*\dagger}$ |
| Model 33 | 3271 | 62685 | 5503 | 540 | 276 | 142$^{*\dagger}$ | 2858 | 213 | 37 | 16$^{*}$ |
| *2 weeks ahead* | | | | | | | | | | |
| Mean | 3045 | 38620 | 6102 | 720 | 219† | 223 | 4031 | 357 | 64 | 24$^{*}$ |
| Median | 3303 | 47609 | 6284 | 737 | 282 | 200 | 3879 | 297 | 64 | 24$^{*}$ |
| Ensemble | 3231 | 44870 | 6220 | 740 | 285 | 224 | 4037 | 359 | 64 | 24$^{*}$ |
| Sym trim | 3105 | 39643 | 6213 | 728 | 223$^{\S}$ | 202 | 3983 | 297 | 63$^{*}$ | 24$^{*}$ |
| Interior trim | 3491 | 32197 | 7897 | 729 | 158$^{*\dagger\S}$ | 224 | 4037 | 359 | 64 | 24$^{*}$ |
| Previous best | 3063 | 40956 | 5797 | 928 | 235 | 204 | 3354$^{*}$ | 321 | 81 | 26 |
| Inv score | 2842$^{\dagger\S}$ | 31725$^{*}$ | 5927$^{\dagger}$ | 713$^{*}$ | 188$^{\dagger\S}$ | 217 | 3898$^{\dagger}$ | 347 | 64 | 24$^{*\dagger}$ |
| Inv score tuning | 2756$^{*\S}$ | 32620 | 5601$^{*\S}$ | 738 | 171$^{\S}$ | 193$^{*}$ | 3651 | 289$^{*}$ | 64 | 24$^{*}$ |
| Model 33 | 3332 | 48704 | 6166 | 856 | 305 | 197 | 3601 | 300 | 67 | 24$^{*}$ |
| *3 weeks ahead* | | | | | | | | | | |
| Mean | 3594 | 43109 | 7178 | 1022 | 257$^{\S}$ | 303 | 5022 | 497 | 100 | 33$^{*}$ |
| Median | 3795 | 52918 | 7100 | 1077 | 318 | 265 | 4769 | 400 | 98$^{*}$ | 34 |
| Ensemble | 3655 | 46027 | 7070 | 1075 | 329 | 263 | 4674 | 397 | 98$^{*}$ | 34 |
| Sym trim | 3777 | 48037 | 7423 | 1041 | 263$^{\S}$ | 270 | 4996 | 399 | 98$^{*}$ | 33$^{*}$ |
| Interior trim | 4503 | 36011 | 10377 | 1050 | 230$^{\S}$ | 305 | 5058 | 500 | 101 | 34 |
| Previous best | 3766 | 53485 | 6604 | 1425 | 344 | 277 | 4129$^{*}$ | 442 | 127 | 37 |
| Inv score | 3349$^{\dagger}$ | 35893 | 6899$^{\dagger}$ | 1012$^{*}$ | 221$^{*\dagger\S}$ | 292 | 4763$^{\dagger}$ | 480 | 100 | 33$^{*\dagger}$ |
| Inv score tuning | 3177$^{*}$ | 39846 | 6090$^{*\S}$ | 1052 | 232$^{\S}$ | 255$^{*}$ | 4378 | 389$^{*}$ | 99 | 33$^{*}$ |
| Model 33 | 3442 | 35365$^{*}$ | 6859 | 1229 | 361 | 257 | 4220 | 403 | 102 | *34* |
| *4 weeks ahead* | | | | | | | | | | |
| Mean | 4158 | 50151 | 8096 | 1373 | 300$^{\S}$ | 395 | 6422 | 642 | 143 | 44 |
| Median | 4580 | 66996 | 8219 | 1471 | 379 | 350 | 6070 | 530 | 140$^{*}$ | 44 |
| Ensemble | 4387 | 56047 | 8235 | 1495 | 393 | 348 | 6016 | 526 | 140$^{*}$ | 45 |
| Sym trim | 4526 | 58825 | 8629 | 1444 | 312$^{\S}$ | 355 | 6387 | 527 | 140$^{*}$ | 44$^{*}$ |
| Interior trim | 5335 | 42720 | 12085 | 1413 | 308 | 398 | 6503 | 647 | 144 | 44 |
| Previous best | 4734 | 70708 | 7799 | 2053 | 470 | 381 | 5654 | 600 | 183 | 50 |
| Inv score | 3799$^{\dagger}$ | 38510 | 7736$^{\dagger}$ | 1345$^{*}$ | 275$^{*\dagger\S}$ | 381 | 6095$^{\dagger}$ | 619 | 143 | 43$^{*\dagger}$ |
| Inv score tuning | 3731$^{*}$ | 50936 | 6690$^{*\S}$ | 1428 | 298$^{\S}$ | 337 | 5617 | 514$^{*}$ | 142 | 44 |
| Model 33 | 3781 | 30433 | 7680 | 1646 | 450 | 328$^{*}$ | 4963$^{*}$ | 522 | 145 | 46 |

$^{*}$For each horizon, best method for each column; $^{\dagger}$score is significantly lower than the mean combination; $^{\S}$ score is significantly lower than the median combination. Lower values are better.
