## Supplemental Table 4 for "Forecasts of weekly incident and cumulative COVID-19 mortality in the United States: A comparison of combining methods"

**S4 Table. For incident COVID-19 mortality, scores for each 13 week period.**

| | Mean interval score | | | | | Mean absolute error | | | | |
|---|---|---|---|---|---|---|---|---|---|---|
| **Method** | **All** | **U.S.** | **High** | **Med** | **Low** | **All** | **U.S.** | **High** | **Med** | **Low** |
| *First period* | | | | | | | | | | |
| Mean | 503 | 5992* | 978 | 166 | 42 | 56 | 1064 | 81 | 23 | 6 |
| Median | 419* | 6297 | 728 | 150* | 34* | 48* | 821 | 73* | 22* | 5* |
| Model 1 | 450 | 7226 | 703* | 199 | 48 | 52 | 772* | 84 | 23 | 6 |
| *Second period* | | | | | | | | | | |
| Mean | 452 | 7683 | 668 | 206* | 57 | 53 | 1004 | 68 | 27* | 9 |
| Median | 536 | 8361 | 791 | 278 | 79 | 55 | 1020 | 70 | 28 | 10 |
| Ensemble | 547 | 8570 | 803 | 286 | 79 | 55 | 1006 | 71 | 28 | 10 |
| Sym trim | 522 | 8756 | 745 | 267 | 71 | 54 | 1020 | 69 | 28 | 9 |
| Exterior trim | 586 | 11146 | 771 | 289 | 77 | NA | NA | NA | NA | NA |
| Interior trim | 468 | 7683 | 717 | 208 | 55 | NA | NA | NA | NA | NA |
| Previous best | 456 | 5649 | 638 | 313 | 112 | 53 | 826 | 74 | 30 | 10 |
| Inv score | 416 | 7147 | 593 | 207 | 53* | 50 | 876 | 66 | 27* | 9 |
| Inv score tuning | 416 | 7147 | 593 | 207 | 53* | 50 | 876 | 66 | 27* | 9 |
| Model 1 | 373* | 6075 | 465* | 251 | 67 | 44* | 593 | 62* | 29 | 9 |
| Model 33 | 463 | 5505* | 718 | 301 | 72 | 46 | 566* | 71 | 28 | 8* |
| *Third period* | | | | | | | | | | |
| Mean | 1148 | 14161 | 2156 | 391 | 132 | 143 | 2874 | 189 | 60 | 20 |
| Median | 1168 | 18290 | 1956 | 405 | 135 | 143 | 2819 | 193 | 59* | 20 |
| Ensemble | 1203 | 17663 | 2066 | 434 | 142 | 143 | 2787 | 194 | 59* | 20 |
| Sym trim | 1216 | 15104 | 2290 | 399 | 141 | 144 | 2819 | 195 | 59* | 20 |
| Exterior trim | 1325 | 18193 | 2413 | 417 | 152 | NA | NA | NA | NA | NA |
| Interior trim | 1204 | 14325 | 2341 | 377 | 122* | NA | NA | NA | NA | NA |
| Previous best | 1307 | 21529 | 1903 | 671 | 158 | 167 | 3436 | 214 | 70 | 26 |
| Inv score | 1029 | 12431 | 1914 | 377* | 125 | 139 | 2685 | 187 | 60 | 20 |
| Inv score tuning | 838* | 11675* | 1343* | 396 | 139 | 128 | 2140 | 183 | 61 | 20 |
| Model 1 | 1155 | 21363 | 1520 | 613 | 142 | 145 | 2857 | 193 | 64 | 18* |
| Model 33 | 1051 | 14483 | 1634 | 530 | 198 | 121* | 1924* | 179* | 60 | 20 |
| *Fourth period* | | | | | | | | | | |
| Mean | 1125 | 7176 | 2125 | 769 | 124 | 99 | 1181 | 183 | 41 | 10 |
| Median | 595 | 6391 | 682 | 646 | 116 | 56 | 953 | 72 | 34* | 9* |
| Ensemble | 594* | 6624 | 668* | 645* | 114* | 57 | 1001 | 71* | 34* | 9* |
| Sym trim | 890 | 6968 | 1499 | 701 | 114* | 85 | 953 | 160 | 35 | 9* |
| Exterior trim | 1078 | 7176 | 1987 | 765 | 124 | NA | NA | NA | NA | NA |
| Interior trim | 1022 | 7552 | 1828 | 739 | 116 | NA | NA | NA | NA | NA |
| Previous best | 856 | 7529 | 1076 | 965 | 133 | 74 | 1315 | 96 | 42 | 10 |
| Inv score | 875 | 7551 | 1389 | 726 | 118 | 96 | 1131 | 178 | 40 | 10 |
| Inv score tuning | 782 | 7768 | 1055 | 758 | 121 | 66 | 994 | 92 | 40 | 10 |
| Model 1 | 638 | 5108* | 828 | 690 | 133 | 55* | 829* | 75 | 36 | 10 |
| Model 33 | 722 | 5782 | 1036 | 686 | 146 | 70 | 1363 | 87 | 37 | 11 |

* For each period, best method for each column. Lower values are better.
