## Supplemental Table 5 for "Forecasts of weekly incident and cumulative COVID-19 mortality in the United States: A comparison of combining methods"

**S5 Table. For cumulative COVID-19 mortality, scores for each 13 week period.**

| | Mean interval score | | | | | Mean absolute error | | | | |
|---|---|---|---|---|---|---|---|---|---|---|
| **Method** | **All** | **U.S.** | **High** | **Med** | **Low** | **All** | **U.S.** | **High** | **Med** | **Low** |
| *First period* | | | | | | | | | | |
| Mean | 1568* | 14137 | 3516* | 301 | 148 | 140* | 1863* | 258 | 46 | 15 |
| Median | 1811 | 14288 | 4268 | 275 | 156 | 144 | 2318 | 250 | 42* | 13* |
| Model 1 | 1686 | 12850* | 3992 | 267* | 141* | 140* | 2034 | 252* | 43* | 14* |
| *Second period* | | | | | | | | | | |
| Mean | 1099 | 15757 | 1813 | 505 | 116 | 126 | 2541 | 162 | 56 | 18 |
| Median | 1296 | 17304 | 2136 | 653 | 156 | 128 | 2508 | 167 | 59 | 19 |
| Ensemble | 1314 | 17665 | 2163 | 658 | 159 | 127 | 2425 | 166 | 59 | 19 |
| Sym trim | 1331 | 23368 | 1990 | 570 | 137 | 127 | 2530 | 163 | 57 | 19 |
| Exterior trim | 1444 | 26815 | 2081 | 613 | 146 | NA | NA | NA | NA | NA |
| Interior trim | 937* | 15757 | 1338* | 486* | 115 | NA | NA | NA | NA | NA |
| Previous best | 1153 | 10366* | 2031 | 680 | 207 | 118 | 1467 | 186 | 68 | 20 |
| Inv score | 943 | 12051 | 1575 | 489 | 110* | 120 | 2313 | 158* | 56* | 18 |
| Inv score tuning | 943 | 12051 | 1575 | 489 | 110* | 120 | 2313 | 158* | 56* | 18 |
| Model 1 | 1282 | 16376 | 2079 | 685 | 192 | 134 | 1576 | 198 | 91 | 28 |
| Model 33 | 1236 | 14458 | 2081 | 701 | 149 | 104* | 1317* | 165 | 60 | 17* |
| *Third period* | | | | | | | | | | |
| Mean | 6747 | 103950 | 13424 | 690 | 409 | 445 | 9141 | 661 | 115 | 49 |
| Median | 7735 | 132486 | 14540 | 790 | 538 | 440 | 8696 | 670 | 115 | 49 |
| Ensemble | 7436 | 115628 | 14588 | 797 | 560 | 435 | 8479 | 669 | 115 | 49 |
| Sym trim | 6952 | 107828 | 13792 | 712 | 418 | 452 | 9276 | 675 | 114 | 48 |
| Exterior trim | 6943 | 107979 | 13733 | 727 | 426 | NA | NA | NA | NA | NA |
| Interior trim | 4374* | 70222* | 8224* | 747 | 278* | NA | NA | NA | NA | NA |
| Previous best | 6833 | 104759 | 13106 | 1231 | 402 | 456 | 8002 | 721 | 150 | 55 |
| Inv score | 6243 | 83841 | 13147 | 689* | 327 | 439 | 8894 | 659 | 114 | 48 |
| Inv score tuning | 6101 | 84170 | 12660 | 741 | 311 | 420 | 8058 | 655 | 110* | 47* |
| Model 1 | 16575 | 359767 | 24754 | 3511 | 1271 | 687 | 15911 | 908 | 188 | 69 |
| Model 33 | 6794 | 101831 | 13186 | 1000 | 607 | 404* | 7123* | 648* | 120 | 49 |
| *Fourth period* | | | | | | | | | | |
| Mean | 2661 | 22735 | 4868 | 1693 | 243 | 235 | 2216 | 479 | 91 | 20 |
| Median | 1848 | 13909* | 3164 | 1443* | 229 | 138* | 1743 | 221 | 78* | 19* |
| Ensemble | 1804* | 14326 | 2999* | 1450 | 225 | 139 | 1882 | 216* | 79 | 20 |
| Sym trim | 2625 | 22735 | 4885 | 1594 | 214* | 138* | 1708* | 220 | 81 | 19* |
| Exterior trim | 2714 | 22735 | 5004 | 1724 | 237 | NA | NA | NA | NA | NA |
| Interior trim | 6939 | 21280 | 18101 | 1638 | 234 | NA | NA | NA | NA | NA |
| Previous best | 2488 | 30259 | 3625 | 1904 | 301 | 174 | 2265 | 269 | 108 | 21 |
| Inv score | 2656 | 24009 | 4791 | 1692 | 228 | 223 | 2173 | 443 | 91 | 20 |
| Inv score tuning | 2163 | 24744 | 3191 | 1729 | 240 | 155 | 2211 | 228 | 95 | 20 |
| Model 1 | 3379 | 18523 | 6319 | 2438 | 488 | 194 | 2656 | 310 | 102 | 26 |
| Model 33 | 2246 | 14769 | 4131 | 1588 | 283 | 179 | 3271 | 250 | 82 | 24 |

[a] For each quarter, best method for each column. Lower values are better.
